# Cannabis and cannabinoids in dermatology: protocol for a systematic review and meta-analysis of quantitative outcomes

**DOI:** 10.1101/2023.03.05.23286815

**Authors:** Pim Sermsaksasithorn, Tanawin Nopsopon, Chatpol Samuthpongtorn, Korn Chotirosniramit, Krit Pongpirul

## Abstract

Following the discovery of various effects on skin function by modifying endocannabinoid systems, multiple preclinical studies have revealed the promise of cannabis and cannabinoids in the treatment of a variety of skin diseases. However, its clinical efficacy is still debated. This systematic review aims to evaluate the therapeutic efficacy of cannabis and cannabinoids in dermatological conditions and diseases. The protocol has been prepared using the Preferred Items for Systematic Review and Meta-analysis Protocols (PRISMA-P) guidelines and has been registered with the International Prospective Register of Systematic Reviews (PROSPERO: CRD42023397189). PubMed, EMBASE, SCOPUS, Cochrane Central Register of Controlled Trials (CENTRAL), and Web of Science will be used for the systemic search. The outcomes will be an improvement in dermatological characteristics, including disease-specific composite scores and objective and subjective outcomes of skin conditions. This systematic review will provide valuable information on dermatological therapy of cannabis and cannabinoids and could be the start of future research and medicinal applications.

## Introduction

Skin is the largest human organ and is essential for organism survival. It is the first line of immunological and physical protection against the external environment, including heat control and retention of hydration [1, 2]. A substantial part of the population is affected by skin diseases that have significant effects on the quality of life of sufferers (3-7).

The Endocannabinoid System (ECS), which includes receptors, ligands, and enzymes, is mainly responsible for maintaining homeostasis and the balance of multiple biological functions in the nervous system and peripheral organs [3-5]. Cannabinoids and active components of Cannabis sativa, which were found to mimic endocannabinoid signaling and influence receptor expression [6, 7], have gained interest as potential treatment for various diseases [3, 5]. The growing legalization of medicinal cannabis and cannabinoids led to the search for medicinal use in clinical practice. The FDA currently approves the use of cannabinoid to alleviate pain and spasticity in multiple sclerosis and the treatment of chemotherapy-induced nausea and vomiting in cancer patients [8-11].

Modulation of the activity of the endocannabinoid system has been shown to influence several types of skin disease, including atopic dermatitis, psoriasis, acne, and skin tumors, according to the etiology of the diseases [12-15]. Furthermore, cannabis has anti-inflammatory and anti-pruritic properties [10, 16-18]. This makes the cannabinoids and active components of Cannabis sativa promising for the therapeutic treatment of the skin disorders mentioned above. Given the insufficient quality clinical studies on this topic, this systematic review aims to examine the efficacy of cannabis and cannabinoids in alleviating dermatological conditions and diseases by highlighting current and historical attempts to collect robust clinical data and identify knowledge gaps.

## Material and methods

The systematic review protocol was registered with the International Prospective Register of Systematic Reviews (PROSPERO) on February 17^th^, 2023 (CRD42023397189). The protocol followed the Preferred Items for Systematic Review and Meta-analysis Protocols (PRISMA-P) guidelines.

### Study Selection

Any randomized controlled trials and observational studies including cross-sectional studies, case-control studies, or cohort studies that investigate the efficacy or effectiveness of any form of medical cannabis or cannabinoid in alleviating dermatological conditions or diseases will be included. Exclusion criteria are (1) in vitro studies, case report, protocol, review article, guideline, editorial, commentary, and letter to editor (2) non-peer-reviewed studies (3) animal studies (4) studies published in non-English. Human participants of all ages will be included. The intervention includes an application of cannabis or cannabinoids in any preparation through any route. Study with placebo control, active control, or even no control will be assessed.

### Study outcome

The efficacy or effectiveness of cannabis and cannabinoids in alleviating dermatological conditions or diseases will be determined based on the followings:

1. Generic outcomes used to assess improvement in skin conditions:
  a. Subjective clinical or patient-reported outcomes such as pruritus score, erythema grade, and quality of life.
  b. Objective evaluation using standard instruments such as transepidermal water loss, lipid analysis, and skin topography evaluation.
2. Disease specific composite scores, mixed subjective and objective outcomes, such as Psoriasis Area Severity Index (PASI) for psoriasis, Eczema Area and Severity Index (EASI), and SCORing Atopic Dermatitis (SCORAD) for atopic dermatitis, etc.

### Search strategy

We will search through five databases: PUBMED, EMBASE, SCOPUS, Cochrane Central Register of Controlled Trials (CENTRAL), and Web of Science. The search strategy constructed by two health information specialists with systematic review experience will combine search terms and subject headings (MeSH) related to ‘cannabis’, ‘cannabaceae’, ‘cannabinoids’, ‘dronabinol’, ‘dermatology’, ‘skin’, and ‘ulcer’ (Supplementary 1).

### Study records

#### Data management

After deriving the studies via the mentioned database, we will import them into Covidence systematic review software, which de-duplicates studies and facilitates study selection [19].

#### Selection process

The titles and abstracts of the identified citations will be evaluated by independent paired reviewers, and initially, the abstracts that do not report the therapeutic effects of cannabis and cannabinoids in dermatological conditions or diseases will be eliminated.

The included studies will then undergo full text review and the final included study will be selected based on all eligibility criteria. Reasons for study exclusion in this step will be recorded. When differences could not be resolved through dialogue, an adjudicator will be brought in to assist. The PRISMA 2020 flow chart will be created to illustrate the workflow.

#### Data collection and management

The data extraction criteria will be refined prior to data collection to ensure consistency among reviewers. The extracted data includes (1) study characteristics including authors, year of publication, study design, journal, contact information, country and funding (2) participant information including mean age, sex, number of participants, type and baseline severity of skin diseases or conditions (3) treatment details including the kind of medical cannabinoid used, additional constituents of intervention products, route of administration and the length of treatment (4) control preparation, route of administration, and duration of application (5) dermatological improvement incoporating results and time points of reported outcome (6) missing data (7) interpretation and discussion (8) all relevant text, tables, and figures. We will contact the corresponding authors of the included studies to obtain incompletely reported data. If no response is received within 14 days, studies will be carried out using the available data.

### Risk of bias

The two reviewers will independently assess the risk of bias. The risk of bias of all randomized controlled studies will be assessed using the Cochrane Risk-of-Bias 2 (RoB2), including the randomization process, allocation concealment, blinding of participants, outcome evaluation, fully addressed outcome data, selective outcome reporting, and other sources of bias. Using ROBINS-1, bias of all nonrandomized controlled studies was evaluated, including bias due to confounding, selection bias, bias in classification of interventions, bias due to deviations from intended interventions, bias due to missing data, bias in measurement of outcomes and bias in selection of the reported result. When a dispute between two reviewers cannot be resolved through dialogue, an adjudicator will be called in to aid.

### Data synthesis

#### Qualitative synthesis

We will qualitatively analyze the studies and their results in accordance with Standard 4.2 and Chapter 4 of Finding What Works in Health Care: Standards for Systematic Review [20]. We will analyze the studies following the study outcomes, discuss the details of each performance of cannabis and cannabinoids in alleviating dermatological conditions or diseases, and evaluate the risk of bias.

#### Quantitative systhesis

If the study includes a control group, we will evaluate the efficacy of medical cannabis or cannabinoids in alleviating skin conditions between case and control, as well as between pre- and post-cannabinoids applications. If the study lacks a control group, we will only compare pre- and post-performance. We will combine the study results for each outcome reported in common by two or more studies using the standard mean difference (SMD) method for continuous outcomes and the relative risk (RR) method for dichotomous outcomes. For continuous outcomes in studies with controls, we will utilize the standard mean difference (SMD) of the difference between the case’s pre- and post-performance and the control’s pre- and post-performance to eliminate baseline heterogeneity between case and control. To compare findings before and after cannabinoid treatment, we will use the standard mean difference (SMD) of outcomes before and after cannabinoid administration. The pooled effect sizes and 95% confidence intervals (CI) will be calculated using random effects models.

For continuous outcomes measured on different scales, in addition to the standard mean difference (SMD) approach for standardization [21], we will attempt to employ the odd ratio method (OR) by specifying the cutoff of the outcome and converting all continuous measures to binary scale, which are ‘improvement’ and ‘no improvement’. We will seek the raw data from the respective authors by contacting them. If we were unable to get the response within 14 days, we will perform the analysis using only the available data.

RevMan 5.4 (The Cochrane Collaboration, The Nordic Cochrane Centre, Copenhagen, Denmark) will be used to perform the meta-analysis.P-value < 0.05 will be considered statistically significant.

#### Assessment of heterogeneity

Heterogeneity will be determined using the Cochran’s Q test [a p-value of 0.10 indicated heterogeneity] and the Higgins’ test [I2] [less than 25%: low heterogeneity, 25–75%: moderate heterogeneity, more than 75%: high heterogeneity] [22].

Sensitivity analysis and Publication bias Sensitivity analyzes will examine redoing the meta-analysis by removing one research at a time to assess the statistical robustness of the primary outcome. The Egger’s regression asymmetry test and funnel plots will be considered to assess publication bias using R version 4.0.1 if the number of identified studies is fewer than 10.

## Discussion

Pathological investigations have suggested that cannabis and cannabinoids may be beneficial in the treatment of dermatological problems. However, there is insufficient agreement on their true therapeutic applicability. Previous scoping studies and systematic reviews may have focused on different types of skin disorders; nevertheless, quantitative meta-analysis and systematic reviews based primarily on skin problems tend to be restricted, particularly when dividing the measured results into subjective, objective and disease-specific composite scores. Reporting the efficacy of the treatment in the aforementioned characteristics can make the findings in this study easier to apply, since this quantitative reporting can be easier to comprehend and reproduce for future extended research and decreases the biased bias of the authors, and because a skin condition is identified in many skin diseases. This systematic review and meta-analysis aims to provide a comprehensive summary of the therapeutic effects of cannabis and cannabinoids on dermatological conditions and diseases. This informs clinicians and patients about the efficacy of cannabis and cannabinoids in skin disorders and identifies information gaps that may lead to the creation of a new alternative therapy for dermatological diseases.

## Data Availability

All data produced in the present study are available upon reasonable request to the authors.

## Data Availability

All data produced in the present study are available upon reasonable request to the authors.

