## Supplementary 1 for "Cannabis and cannabinoids in dermatology: protocol for a systematic review and meta-analysis of quantitative outcomes"

Concept 1: Cannabinoid
Concept 2: Dermatology

| Set # | Pubmed (until 9 Nov 2022) |
|---|---|
| 1 Cannabinoids | "cannabis"[MeSH Terms] OR cannabis[tiab] OR canabis[tiab] OR "cannabaceae"[MeSH Terms:noexp] OR cannabaceae[tiab] OR hemp*[tiab] OR marijuana[tiab] OR marihuana[tiab] OR ganja*[tiab] OR hash[tiab] OR hashish[tiab] OR bhang[tiab] OR skunk[tiab] OR sinsemilla[tiab] OR charas[tiab] OR weed*[tiab] OR "cannabinoids"[MeSH Terms] OR cannabinoid*[tiab] OR canabinoid*[tiab] OR cannabidiol*[tiab] OR canabidiol*[tiab] OR cannabinol[tiab] OR cannador[tiab] OR eucannabinolide[tiab] OR "8001-45-4"[tiab] OR "8063-14-7"[tiab] OR "38458-58-1"[tiab] OR "dronabinol"[MeSH Terms] OR dronabinol*[tiab] OR marinol[tiab] OR deltanyne[tiab] OR ea1477[tiab] OR "ea-1477"[tiab] OR tetranabinex[tiab] OR qcd84924[tiab] OR "qcd-84924"[tiab] OR "7663-50-5"[tiab] OR nabidiolex[tiab] OR "13956-29-1"[tiab] OR "nabilone"[Supplementary Concept] OR nabilone[tiab] OR cesamet*[tiab] OR cpd109514[tiab] OR "cpd-109514"[tiab] OR lilly109514[tiab] OR "lilly-109514"[tiab] OR "51022-71-0"[tiab] OR "HU 211"[Supplementary Concept] OR "HU 211"[tiab] OR "HU-211"[tiab] OR hu211[tiab] OR "HU 210"[tiab] OR "HU-210"[tiab] OR hu210[tiab] OR dexanabinol[tiab] OR "112924-45-5"[tiab] OR "tetrahydrocannabinol-cannabidiol combination"[Supplementary Concept] OR "tetrahydrocannabinol-cannabidiol combination"[tiab] OR nabiximol*[tiab] OR sativex[tiab] OR gw1000[tiab] OR "gw-1000"[tiab] OR sab378[tiab] OR "sab-378"[tiab] OR "56575-23-6"[tiab] OR tetrahydrocannabinol*[tiab] OR "tetra-hydrocannabinol"[tiab] OR 9tetrahydrocannabinol[tiab] OR "delta3-THC"[tiab] OR "delta-3-THC"[tiab] OR "delta-3-tetrahydrocannabinol"[tiab] OR sp104[tiab] OR "sp-104"[tiab] OR "1972-08-3"[tiab] OR "delta9-THC"[tiab] OR "delta-9-THC"[tiab] OR "delta-9-tetrahydrocannabinol"[tiab] OR "5957-75-5"[tiab] OR cannabichromene[tiab] OR "521-35-7"[tiab] OR "8-THC"[tiab] OR tetrahydrocannabivarin[tiab] OR anandamide[tiab] OR "n-arachidonoylethanolamine"[tiab] OR nantradol[tiab] OR cp44001[tiab] OR "cp-44001"[tiab] OR cp440011[tiab] OR "cp-44001-1"[tiab] OR "cp44001-1"[tiab] OR "72028-54-7"[tiab] OR endocannabinoid*[tiab] OR phytocannabinoid*[tiab] OR sydros[tiab] OR indica[tiab] OR THC[tiab] OR CBD[tiab] OR AEA[tiab] |
| 2 Dermatology | "dermatology"[MeSH Terms] OR dermatol*[tiab] OR "skin"[MeSH Terms] OR skin[tiab] OR cutaneous[tiab] OR wound*[tiab] OR "ulcer"[MeSH Terms] OR ulcer[tiab] |
| 3 | #1 AND #2 |
| 4 | animals[MeSH Terms] NOT humans[MeSH Terms] |
| 5 | #3 NOT #4 |
| 6 | english[lang] |
| 7 | #5 AND #6 |
| 8 | 1945/1/01:2022/11/09[dp] |
| 9 | #7 AND #8 |

| Set # | Embase (until 9 Nov 2022) |
|---|---|
| 1<br>Cannabinoids | 'cannabis'/exp OR 'cannabis':ti,ab OR 'canabis':ti,ab OR 'cannabaceae'/de OR 'cannabaceae':ti,ab OR 'hemp*':ti,ab OR 'marijuana':ti,ab OR 'marihuana':ti,ab OR 'ganja*':ti,ab OR 'hash':ti,ab OR 'hashish':ti,ab OR 'bhang':ti,ab OR 'skunk':ti,ab OR 'sinsemilla':ti,ab OR 'charas':ti,ab OR 'weed*':ti,ab OR 'cannabinoid'/exp OR 'cannabinoid*':ti,ab OR 'canabinoid*':ti,ab OR 'cannabidiol*':ti,ab OR 'canabidiol*':ti,ab OR 'cannabinol':ti,ab OR 'cannador':ti,ab OR 'eucannabinolide':ti,ab OR '8001-45-4':ti,ab OR '8063-14-7':ti,ab OR '38458-58-1':ti,ab OR 'dronabinol'/exp OR 'dronabinol*':ti,ab OR 'marinol':ti,ab OR 'deltanyne':ti,ab OR 'ea1477':ti,ab OR 'ea-1477':ti,ab OR 'tetranabinex':ti,ab OR 'qcd84924':ti,ab OR 'qcd-84924':ti,ab OR '7663-50-5':ti,ab OR 'nabidiolex':ti,ab OR '13956-29-1':ti,ab OR 'nabilone'/exp OR 'nabilone':ti,ab OR 'cesamet*':ti,ab OR 'cpd109514':ti,ab OR 'cpd-109514':ti,ab OR 'lilly109514':ti,ab OR 'lilly-109514':ti,ab OR '51022-71-0':ti,ab OR 'HU 211':ti,ab OR 'HU-211':ti,ab OR 'hu211':ti,ab OR 'HU 210':ti,ab OR 'HU-210':ti,ab OR 'hu210':ti,ab OR 'dexanabinol'/exp OR 'dexanabinol':ti,ab OR '112924-45-5':ti,ab OR 'tetrahydrocannabinol-cannabidiol combination':ti,ab OR 'nabiximol*':ti,ab OR 'sativex':ti,ab OR 'gw1000':ti,ab OR 'gw-1000':ti,ab OR 'sab378':ti,ab OR 'sab-378':ti,ab OR '56575-23-6':ti,ab OR 'tetrahydrocannabinol*':ti,ab OR 'tetra-hydrocannabinol':ti,ab OR '9tetrahydrocannabinol':ti,ab OR 'delta3-THC':ti,ab OR 'delta-3-THC':ti,ab OR 'delta-3-tetrahydrocannabinol':ti,ab OR 'sp104':ti,ab OR 'sp-104':ti,ab OR '1972-08-3':ti,ab OR 'delta9-THC':ti,ab OR 'delta-9-THC':ti,ab OR 'delta-9-tetrahydrocannabinol':ti,ab OR '5957-75-5':ti,ab OR 'cannabichromene':ti,ab OR '521-35-7':ti,ab OR '8-THC':ti,ab OR 'tetrahydrocannabivarin':ti,ab OR 'anandamide':ti,ab OR 'n-arachidonoylethanolamine':ti,ab OR 'nantradol':ti,ab OR 'cp44001':ti,ab OR 'cp-44001':ti,ab OR 'cp440011':ti,ab OR 'cp-44001-1':ti,ab OR 'cp44001-1':ti,ab OR '72028-54-7':ti,ab OR 'endocannabinoid*':ti,ab OR 'phytocannabinoid*':ti,ab OR 'sydros':ti,ab OR 'indica':ti,ab OR 'THC':ti,ab OR 'CBD':ti,ab OR 'AEA':ti,ab |
| 2<br>Dermatology | 'dermatology'/exp OR 'dermatol*':ti,ab OR 'skin'/exp OR 'skin':ti,ab OR 'cutaneous':ti,ab OR 'wound*':ti,ab OR 'ulcer'/exp OR 'ulcer':ti,ab |
| 3 | #1 AND #2 |
| 4 | [animals]/lim NOT [humans]/lim |
| 5 | #3 NOT #4 |
| 6 | english:la |
| 7 | #5 AND #6 |
| 8 | [09-11-2022]/sd |
| 9 | #7 AND #8 |

| Set # | Scopus (until Nov 2022) |
|---|---|
| 1 Cannabinoids | TITLE-ABS-KEY(cannabis OR cannabis OR cannabaceae OR hemp* OR marijuana OR marihuana OR ganja* OR hash OR hashish OR bhang OR skunk OR sinsemilla OR charas OR weed* OR cannabinoid* OR canabinoid* OR cannabidiol* OR canabidiol* OR cannabinol OR cannador OR eucannabinolide OR "8001-45-4" OR "8063-14-7" OR "38458-58-1" OR dronabinol* OR marinol OR deltanyne OR ea1477 OR "ea-1477" OR tetranabinex OR qcd84924 OR "qcd-84924" OR "7663-50-5" OR nabidiolex OR "13956-29-1" OR nabilone OR cesamet* OR cpd109514 OR "cpd-109514" OR lilly109514 OR "lilly-109514" OR "51022-71-0" OR "HU 211" OR "HU-211" OR hu211 OR "HU 210" OR "HU-210" OR hu210 OR dexanabinol OR "112924-45-5" OR "tetrahydrocannabinol-cannabidiol combination" OR nabiximol* OR sativex OR gw1000 OR "gw-1000" OR sab378 OR "sab-378" OR "56575-23-6" OR tetrahydrocannabinol* OR "tetra-hydrocannabinol" OR 9tetrahydrocannabinol OR "delta3-THC" OR "delta-3-THC" OR "delta-3-tetrahydrocannabinol" OR sp104 OR "sp-104" OR "1972-08-3" OR "delta9-THC" OR "delta-9-THC" OR "delta-9-tetrahydrocannabinol" OR "5957-75-5" OR cannabichromene OR "521-35-7" OR "8-THC" OR tetrahydrocannabivarin OR anandamide OR "n-arachidonoylethanolamine" OR nantradol OR cp44001 OR "cp-44001" OR cp440011 OR "cp-44001-1" OR "cp44001-1" OR "72028-54-7" OR endocannabinoid* OR phytocannabinoid* OR sydros OR indica OR THC OR CBD OR AEA) |
| 2 Dermatology | TITLE-ABS-KEY(dermatol* OR skin OR cutaneous OR wound* OR ulcer) |
| 3 | #1 AND #2 |
| 4 | ALL(animals AND NOT humans) |
| 5 | #3 AND NOT #4 |
| 6 | LANGUAGE(english) |
| 7 | #5 AND #6 |
| 8 | PUBYEAR BEF 2023 |
| 9 | PUBDATETXT(December 2022) |
| 10 | #8 AND NOT #9 |
| 11 | #7 AND #10 |

| Set # | Web of Science (until Dec 2022) |
|---|---|
| 1<br>Cannabinoids | TS=(cannabis OR cannabis OR cannabaceae OR hemp* OR marijuana OR marihuana OR ganja* OR hash OR hashish OR bhang OR skunk OR sinsemilla OR charas OR weed* OR cannabinoid* OR canabinoid* OR cannabidiol* OR canabidiol* OR cannabinol OR cannador OR eucannabinolide OR "8001-45-4" OR "8063-14-7" OR "38458-58-1" OR dronabinol* OR marinol OR deltanyne OR ea1477 OR "ea-1477" OR tetranabinex OR qcd84924 OR "qcd-84924" OR "7663-50-5" OR nabidiolex OR "13956-29-1" OR nabilone OR cesamet* OR cpd109514 OR "cpd-109514" OR lilly109514 OR "lilly-109514" OR "51022-71-0" OR "HU 211" OR "HU-211" OR hu211 OR "HU 210" OR "HU-210" OR hu210 OR dexanabinol OR "112924-45-5" OR "tetrahydrocannabinol-cannabidiol combination" OR nabiximol* OR sativex OR gw1000 OR "gw-1000" OR sab378 OR "sab-378" OR "56575-23-6" OR tetrahydrocannabinol* OR "tetra-hydrocannabinol" OR 9tetrahydrocannabinol OR "delta3-THC" OR "delta-3-THC" OR "delta-3-tetrahydrocannabinol" OR sp104 OR "sp-104" OR "1972-08-3" OR "delta9-THC" OR "delta-9-THC" OR "delta-9-tetrahydrocannabinol" OR "5957-75-5" OR cannabichromene OR "521-35-7" OR "8-THC" OR tetrahydrocannabivarin OR anandamide OR "n-arachidonoylethanolamine" OR nantradol OR cp44001 OR "cp-44001" OR cp440011 OR "cp-44001-1" OR "cp44001-1" OR "72028-54-7" OR endocannabinoid* OR phytocannabinoid* OR sydros OR indica OR THC OR CBD OR AEA) |
| 2<br>Dermatology | TS=(dermatol* OR skin OR cutaneous OR wound* OR ulcer) |
| 3 | #1 AND #2 |
| 4 | ALL=(animal NOT human) |
| 5 | #3 NOT #4 |
| 6 | LA=(English) |
| 7 | #5 AND #6 |
| 8 | PY=2023 |
| 9 | #7 NOT #8 |

| Set # | CENTRAL (until Nov 2022) |
|---|---|
| 1 Cannabinoids | [mh "cannabis"] OR cannabis:ti,ab,kw OR canabis:ti,ab,kw OR [mh "cannabaceae"] OR cannabaceae:ti,ab,kw OR hemp*:ti,ab,kw OR marijuana:ti,ab,kw OR marihuana:ti,ab,kw OR ganja*:ti,ab,kw OR hash:ti,ab,kw OR hashish:ti,ab,kw OR bhang:ti,ab,kw OR skunk:ti,ab,kw OR sinsemilla:ti,ab,kw OR charas:ti,ab,kw OR weed*:ti,ab,kw OR [mh "cannabinoids"] OR cannabinoid*:ti,ab,kw OR canabinoid*:ti,ab,kw OR cannabidiol*:ti,ab,kw OR canabidiol*:ti,ab,kw OR cannabinol:ti,ab,kw OR cannador:ti,ab,kw OR eucannabinolide:ti,ab,kw OR "8001-45-4":ti,ab,kw OR "8063-14-7":ti,ab,kw OR "38458-58-1":ti,ab,kw OR [mh "dronabinol"] OR dronabinol*:ti,ab,kw OR marinol:ti,ab,kw OR deltanyne:ti,ab,kw OR ea1477:ti,ab,kw OR "ea-1477":ti,ab,kw OR tetranabinex:ti,ab,kw OR qcd84924:ti,ab,kw OR "qcd-84924":ti,ab,kw OR "7663-50-5":ti,ab,kw OR nabidiolex:ti,ab,kw OR "13956-29-1":ti,ab,kw OR nabilone:ti,ab,kw OR cesamet*:ti,ab,kw OR cpd109514:ti,ab,kw OR "cpd-109514":ti,ab,kw OR lilly109514:ti,ab,kw OR "lilly-109514":ti,ab,kw OR "51022-71-0":ti,ab,kw OR "HU 211":ti,ab,kw OR "HU-211":ti,ab,kw OR hu211:ti,ab,kw OR "HU 210":ti,ab,kw OR "HU-210":ti,ab,kw OR hu210:ti,ab,kw OR dexanabinol:ti,ab,kw OR "112924-45-5":ti,ab,kw OR "tetrahydrocannabinol-cannabidiol combination":ti,ab,kw OR nabiximol*:ti,ab,kw OR sativex:ti,ab,kw OR gw1000:ti,ab,kw OR "gw-1000":ti,ab,kw OR sab378:ti,ab,kw OR "sab-378":ti,ab,kw OR "56575-23-6":ti,ab,kw OR tetrahydrocannabinol*:ti,ab,kw OR "tetra-hydrocannabinol":ti,ab,kw OR 9tetrahydrocannabinol:ti,ab,kw OR "delta3-THC":ti,ab,kw OR "delta-3-THC":ti,ab,kw OR "delta-3-tetrahydrocannabinol":ti,ab,kw OR sp104:ti,ab,kw OR "sp-104":ti,ab,kw OR "1972-08-3":ti,ab,kw OR "delta9-THC":ti,ab,kw OR "delta-9-THC":ti,ab,kw OR "delta-9-tetrahydrocannabinol":ti,ab,kw OR "5957-75-5":ti,ab,kw OR cannabichromene:ti,ab,kw OR "521-35-7":ti,ab,kw OR "8-THC":ti,ab,kw OR tetrahydrocannabivarin:ti,ab,kw OR anandamide:ti,ab,kw OR "n-arachidonoylethanolamine":ti,ab,kw OR nantradol:ti,ab,kw OR cp44001:ti,ab,kw OR "cp-44001":ti,ab,kw OR cp440011:ti,ab,kw OR "cp-44001-1":ti,ab,kw OR "cp44001-1":ti,ab,kw OR "72028-54-7":ti,ab,kw OR endocannabinoid*:ti,ab,kw OR phytocannabinoid*:ti,ab,kw OR sydros:ti,ab,kw OR indica:ti,ab,kw OR THC:ti,ab,kw OR CBD:ti,ab,kw OR AEA:ti,ab,kw |
| 2 Dermatology | [mh "dermatology"] OR dermatol*:ti,ab,kw OR [mh "skin"] OR skin:ti,ab,kw OR cutaneous:ti,ab,kw OR wound*:ti,ab,kw OR [mh "ulcer"] OR ulcer:ti,ab,kw |
| 3 | #1 AND #2 |
| 4 | Limit to Jan 1996-Nov 2022 |
